# Demographic differences in US adult intentions to receive a potential coronavirus vaccine and implications for ongoing study

**DOI:** 10.1101/2020.09.07.20190058

**Authors:** Richard M. Carpiano

## Abstract

**Background:** The coronavirus pandemic’s public health and economic impacts have led to much hope in the US regarding the prospect of a safe, effective vaccine to either prevent infection or minimize symptoms and reduce mortality risk. However, recent US polls indicate a concerning level of hesitancy that will likely lead to suboptimal uptake if such a vaccine becomes available. This study investigated demographic differences regarding US adults’ intent, uncertainty, and refusal to receive a potential coronavirus vaccine and specific reasons for intention to receive it.

**Methods and findings:** Multivariable analysis of Associated Press (AP)-NORC Center for Public Affairs Research cross-sectional survey data collected in May 2020 from a US nationally representative panel of adults (n = 1000). Respondents were asked if they would receive a coronavirus vaccine (yes, unsure, no). Among those answering yes, the specific reasons were: to protect self, family, and community; chronic health condition; and having a doctor who recommends vaccines. Multinomial logistic regression models indicated numerous subgroup differences between participants who indicating (1) uncertainty versus refusal, (2) intent versus refusal, and (3) intent versus uncertainty, with the highest number of significant differences observed in the third comparison. Overall, higher likelihood of intention to receive the vaccine versus uncertainty and refusal were mostly observed among respondents with a college education or greater, White, non-Hispanic racial-ethnic identity, ages 60 or older, and more liberal (versus conservative) ideology. Despite variation in endorsement across the five reasons for wanting to receive the vaccine, subgroup differences were fairly consistent across these specific reasons when comparing respondents endorsing such intentions versus, respectively, refusal and uncertainty in separate analyses.

**Conclusions:** These findings suggest that the approval of a vaccine will potentially face problems with overall uptake due to uncertainty or refusal and contribute to creating significant demographic disparities in COVID-19 morbidity and mortality risk. Ongoing assessment of such attitudes are needed as Phase III trials proceed, but the findings highlight need for measuring uncertainty and its underlying reasons, as well as multiple types of education and outreach efforts for those who are uncertain as well as avoidant.

## Introduction

The public health and economic impacts of the coronavirus pandemic have led to much hope by US policymakers, news media, and the general public about the prospect of a safe, effective vaccine to either prevent infection or minimize symptoms and reduce mortality risk. However, recent US polls indicate a concerning level of hesitancy that will likely lead to suboptimal uptake if such a vaccine becomes available. (e.g., 1,2) Nevertheless, many polls only offer limited information via reporting findings in aggregate or, at best, broken down into crosstabulations according to select demographic subgroups (versus multivariable models that adjust for confounding factors and incorporate a fuller range of the data for deductive or inductive analyses).

Furthermore, when asking participants if they would be willing to get a coronavirus vaccine, many surveys only provide or report on two options—yes or no—excluding an important “uncertain/unsure” category that aids in more comprehensively assessing the breadth and prevalence of hesitancy attitudes. (1,3) This omission is significant, as vaccine hesitancy is a complex phenomenon that encompasses a spectrum of vaccination decision-making, including degrees of uncertainty and complete refusal. (4)

In addition to these methodological issues, several substantive factors unique to a coronavirus vaccine further complicate understanding public hesitancy towards it. First, any public concern, uncertainty, or interest in its receipt centers on a vaccine that is, as of the time of this writing, hypothetical versus an existing product with a known safety and effectiveness profile like all vaccines currently recommended for children and adults. Second, the development of a coronavirus vaccine has been extensively hyped and financially supported by the US Federal Government’s “Operation Warp Speed” initiative, which aims to significantly expedite the development, approval, and production of a vaccine. Promotion of this initiative to the news media and public by US President Donald Trump and his administration during an election year and amidst a pandemic that has witnessed the politicizing of public health and scientific expertise have raised concerns that this expedited process may lead to the approval of an insufficiently evaluated and potentially unsafe and/or inadequately effective vaccine. (5,6) Lastly, though several vaccine candidates exist in different phases of development and evaluation, their safety and effectiveness are already being challenged by anti-vaccine and COVID-19 science denialism disinformation regarding a coronavirus vaccine and other public health measures that have been spreading online for months. (7)

Motivated by these concerns and issues, the present study aims to contribute to current understanding about potential hesitancy towards a COVID-19 vaccine by evaluating demographic differences in US adults’ intentions to receive a potential coronavirus vaccine and the different motivations for such intentions. Existing evidence on adult vaccination provides much reason to be concerned not simply about overall hesitancy for a coronavirus vaccine, but also how such hesitancy may be more prevalent in specific demographic subgroups. These include racial-ethnic and lower socioeconomic status (SES) groups with lower adult vaccination uptake rates and histories of marginalization and surveillance that contribute to their distrust of government, the health care profession, and medical treatments. (8,9) Such hesitancy could risk exacerbating existing disparities in COVID-19 cases and mortality—especially among African Americans, Latinos, and other racial-ethnic groups as well as lower SES groups. (10,11)

Beyond just race-ethnicity and SES, there are empirical and theoretical reasons to conjecture that other demographic groups may be more likely to receive the vaccine due to their COVID-19 risk profiles: older adults, who experience the highest hospitalization and mortality rates (12); men, who experience a higher COVID-19 mortality rate than women (13); married or cohabitating individuals wanting to protect one’s partner and self; persons who have had a relative or close friend diagnosed with COVID-19; employed individuals, whose occupational conditions may necessitate a vaccine to reduce their exposure risk; and geographic locations, like cities and regions of the country where COVID-19 has been more prevalent. Lastly, given the extent to which scientific information and expertise has been politicized and contributed to most COVID-19 related attitudes and behaviors being divided by political ideology (14), one can hypothesize that persons holding liberal or moderate political beliefs will be more likely to express intent to receive the vaccine than will their more politically conservative counterparts, even after controlling for other demographics.

In addition to examining differences in intention to receive a potential COVID-19 vaccine, it is also important to understand potential *motivations* underlying people’s intentions to vaccinate and how people holding specific motivations might differ from others who are either unsure or have no intention. This study examines five prominent motivations. In addition to self-interested, personal protection reasons, vaccination intentions include the prosocial motivations of protecting one’s family and community (15) as well whether one’s physician or health care provider recommends vaccinations (4). Likewise, having a chronic health condition may be another significant motivation to receive the vaccine due to such conditions being risk factors for severe COVID-19 illness and mortality. (16) This reason may be particularly salient for older adults and persons of lower socioeconomic status and racial-ethnic minority status—all groups that experience disproportionate rates of chronic conditions identified as COVID-19 risk factors. Together, these five intentions may be useful for designing eventual vaccine messaging and campaigns to promote uptake.

## Methods

The data are from the May 14-18, 2020 Associated Press (AP)-NORC Center for Public Affairs Research poll, publicly accessible at https://apnorc.org/projects/expectations-for-acovid-19-vaccine/ and determined by the University of California, Riverside Institutional Review Board to be exempt from review. Stata code for this analysis is available from the author for transparency and reproducibility. While data collection procedures are detailed in-depth elsewhere (17), briefly, data were collected using the AmeriSpeak Omnibus®, a monthly survey using NORC’s probability-based panel designed to be representative of the U.S. household population. Adults ages 18 and older from the 50 states and the District of Columbia were randomly drawn from the panel and 1,056 completed the survey via the web and telephone, with interviews conducted in English and Spanish. The analytic sample for this study consisted of 1000 respondents who had complete information for all study variables.

### Measures

Intention to receive a coronavirus vaccine was assessed by asking respondents, “If a vaccine against the coronavirus becomes available, do you plan to get vaccinated, or not? The response options were “Yes,” “No,” and “Not Sure.”

Respondents who answered “Yes” to the preceding item were then asked eight questions about their specific reasons for getting the vaccine. The present study focuses on five of these specific reasons: I want to protect my (1) family, (2) my community, and (3) myself; (4) I have a chronic health condition; and (5) My doctor recommends vaccines. The remaining three reasons—regarding avoiding getting sick, enabling feelings of safety around others, and the need for most people to vaccinate to return to normal—were analyzed, but not reported here due to their findings being very similar to those for the personal and community protection reasons above. Nevertheless, they provided useful corroborating evidence about the validity of the personal and community items reported here.

Because these reason-specific items were only asked among respondents who answered “yes” to the initial vaccine item, they were each recoded into four-category variables to include the entire study sample: (1) Yes for that specific reason (i.e. respondent planned to get the vaccine for the reason specified in the question); (2) Yes, but not for that reason specified in the question; (3) No (i.e. respondent indicated in the original intention item that they had no plans to get the vaccine); and (4) Unsure (respondent indicated in the original item that they were unsure whether they would get the vaccine).

The independent variables include a range of socioeconomic (education, household income), demographic (race-ethnic identity, age, gender/sex, political ideology, self/family/friend having experienced COVID-19, employment status), and geographic factors (urbanicity and census region of residence) selected based on prior research and recent polling reports on COVID-19-related topics. Table 1 details these variables and their descriptive statistics overall and across specific categories of the dependent variables.

### Analyses

Due to the categorical scale of the dependent variables, I estimated adjusted associations between respondent characteristics and vaccination intention (yes, unsure, no) using multivariable multinomial logistic regression models. Using Stata 16.1, all analyses utilized the svyset feature to incorporate the probability weights. Reported associations [adjusted relative risk ratios (RRRs) and 95% confidence intervals] (18) in the Results section are those observed to be statistically significant at a *p*-value < 0.05.

## Results

### Descriptive Statistics

Figure 1 shows that a substantial percentage of respondents are hesitant about receiving a potential coronavirus vaccine: nearly 20% expressed no plan to get it and approximately 30% were unsure. Among the 50.1% who answered “Yes,” the most common reasons were to protect one’s self (93.3%), family (88.6%), and community (78.0%), while intent due to a chronic health condition (36.2%) or what one’s doctor might endorse (45.3%) were less prevalent.

**Fig. 1:**
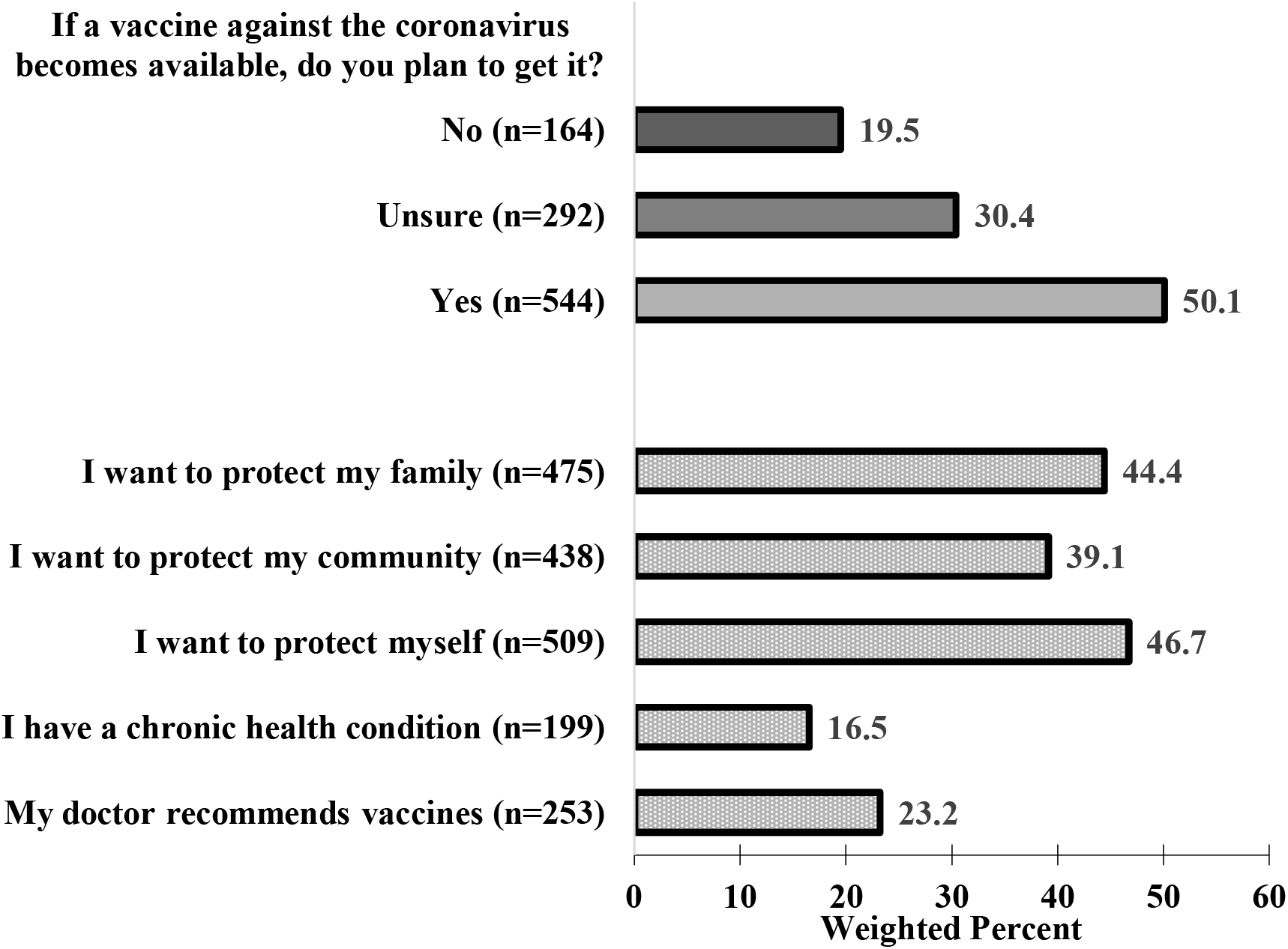
Distribution of the vaccine intention responses (n = 1000). **NOTE:** Reported n’s in parentheses are unweighted. Though specific reasons were only asked of those respondents who answered “Yes” to whether they plan to get a coronavirus vaccine, the weighted percentage reported for each reason is based on the entire sample (n = 1000), which includes all respondents offering that reason as well as those reporting that they (a) plan to get the vaccine, but for a different reason than the one specified reason, (b) do no plan to get a coronavirus vaccine, and (c) are unsure about getting the vaccine.

Table 1 reports the weighted percentages for each independent variable across the overall sample and for the three categories of vaccine intent.

**Table 1:**
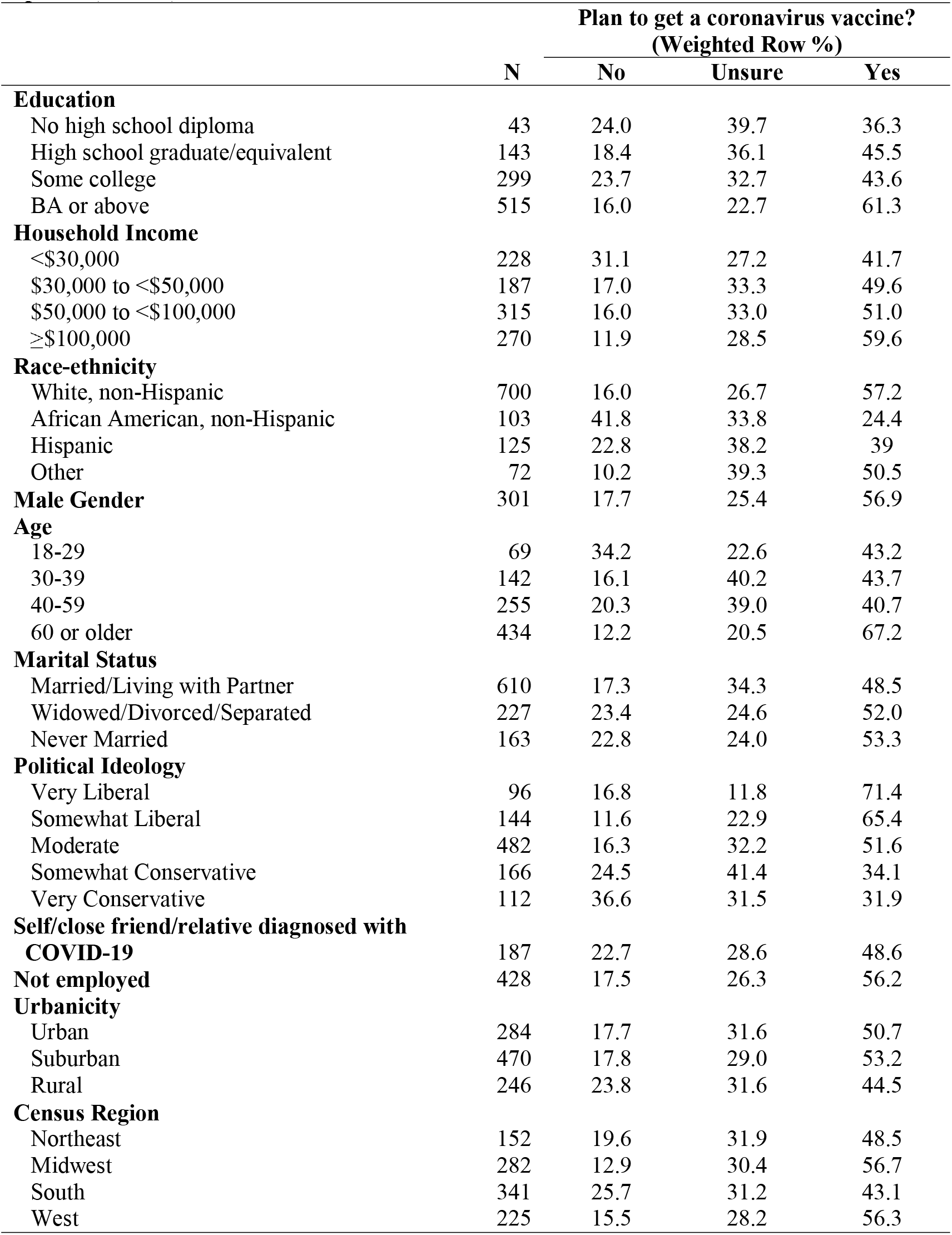
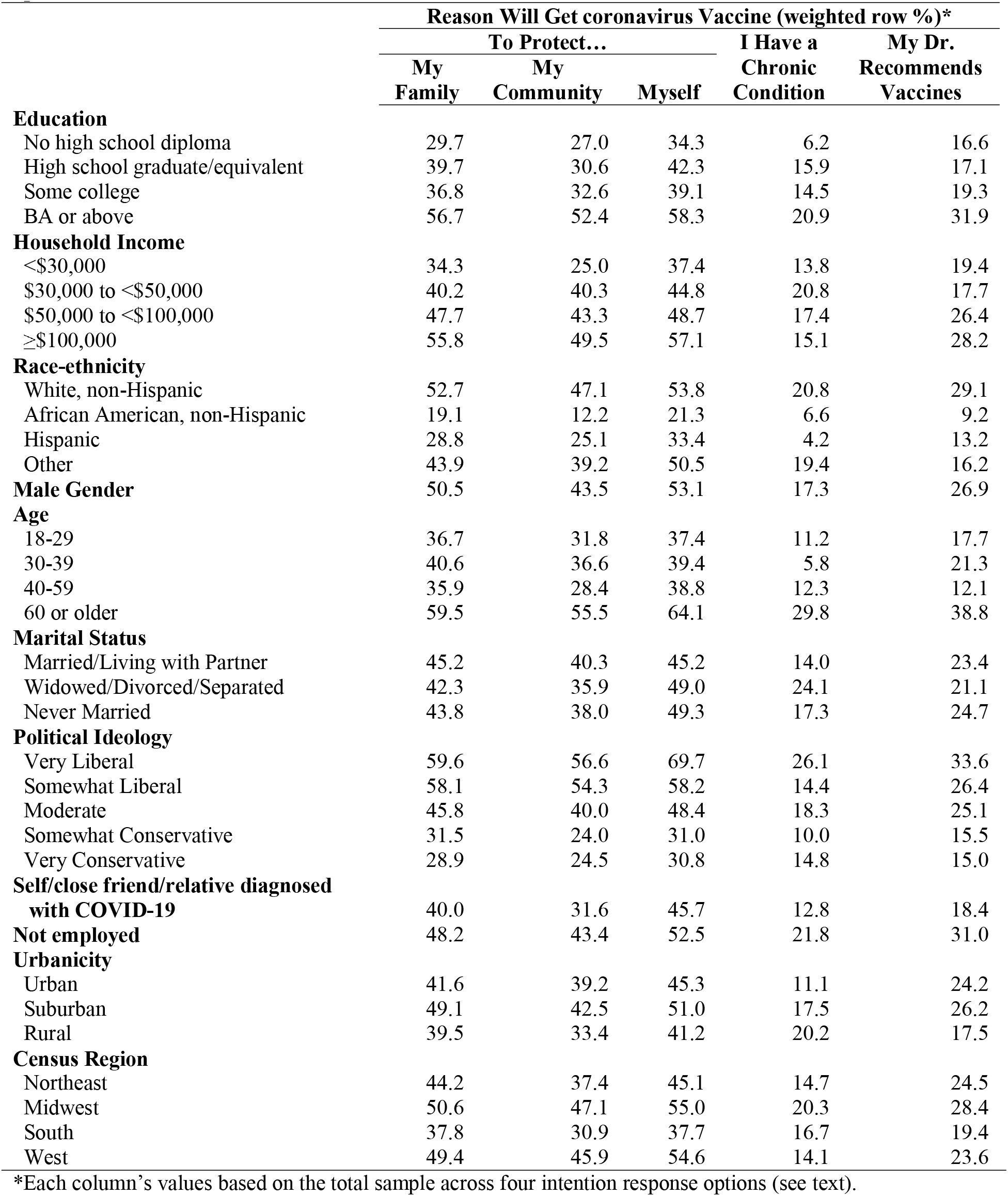
Descriptive statistics for independent variables across coronavirus vaccine intention options (n = 1000).

### If A Coronavirus Vaccine Becomes Available, Who Will Want to Get It?

Table 2 shows RRRs from multinomial logistic regression models comparing the three vaccine intention response categories—No, Unsure, and Yes—against each other. Several patterns are evident, especially with respect to SES, race-ethnicity, age, and political beliefs.

**Table 2:** Adjusted relative risk ratios from multinomial logistic models estimating associations between demographic characteristics and intentions to receive a coronavirus vaccine (n = 1000).

|  | If a Vaccine Becomes Available, Do You Plan to Get it? |  |  |
| --- | --- | --- | --- |
|  | Unsure vs. No | Yes vs. No | Yes vs. Unsure |
| **Education** (referent=BA or higher) |  |  |  |
| No High School diploma | 2.97<br>(0.95, 9.25) | 1.02<br>(0.35, 2.99) | **0.34\***<br>(0.13, 0.88) |
| High School graduate/equivalent | **3.18\*\***<br>(1.55, 6.53) | 1.66<br>(0.82, 3.36) | **0.52\***<br>(0.28, 0.96) |
| Some college | 1.40<br>(0.78, 2.52) | 0.75<br>(0.44, 1.30) | **0.54\*\***<br>(0.34, 0.86) |
| **Household Income** (referent=\$100,000 or more) | | | |
| <\$30,000 | **0.33\***<br>(0.14, 0.78) | **0.24\*\*\***<br>(0.11, 0.53) | 0.72<br>(0.35, 1.48) |
| \$30,000 to <\$50,000 | 0.62<br>(0.26, 1.46) | 0.54<br>(0.24, 1.21) | 0.87<br>(0.46, 1.64) |
| \$50,000 to <\$100,000 | 0.67<br>(0.31, 1.44) | 0.57<br>(0.28, 1.14) | 0.85<br>(0.50, 1.43) |
| **Race-ethnicity** (referent=White, non-Hispanic) |  |  |  |
| African American, non-Hispanic | **0.39\***<br>(0.18, 0.85) | **0.12\*\*\***<br>(0.05, 0.28) | **0.30\*\***<br>(0.13, 0.70) |
| Hispanic | 1.30<br>(0.56, 3.01) | 0.59<br>(0.26, 1.36) | **0.46\***<br>(0.24, 0.89) |
| Other | 2.23<br>(0.77, 6.51) | 1.04<br>(0.36, 3.05) | **0.47\***<br>(0.22, 0.98) |
| **Male** (referent=Female) | 0.77<br>(0.45, 1.33) | 1.60<br>(0.94, 2.70) | **2.07\*\***<br>(1.33, 3.24) |
| **Age** (referent=60 or older) |  |  |  |
| 18-29 | **0.21\*\***<br>(0.08, 0.54) | **0.14\*\*\***<br>(0.06, 0.36) | 0.69<br>(0.30, 1.58) |
| 30-39 | 0.98<br>(0.39, 2.45) | **0.33\***<br>(0.14, 0.78) | **0.34\*\***<br>(0.17, 0.68) |
| 40-59 | 0.84<br>(0.41, 1.72) | **0.33\*\***<br>(0.17, 0.65) | **0.40\*\*\***<br>(0.23, 0.68) |
| **Marital Status** (referent=Married/Living with Partner) |  |  |  |
| Widowed/Divorced/Separated | 0.58<br>(0.27, 1.22) | 0.88<br>(0.44, 1.77) | 1.53<br>(0.83, 2.84) |
| Never Married | 1.35<br>(0.60, 3.06) | **2.48\***<br>(1.15, 5.36) | 1.83<br>(0.97, 3.45) |
| **Political Ideology** (referent=Very Liberal) |  |  |  |
| Somewhat Liberal | 3.51<br>(0.95, 13.0) | 1.88<br>(0.59, 5.95) | 0.54<br>(0.20, 1.42) |
| Moderate | **3.34\***<br>(1.14, 9.81) | 0.79<br>(0.31, 2.04) | **0.24\*\*\***<br>(0.11, 0.53) |
| Somewhat Conservative | 2.09<br>(0.69, 6.37) | **0.21\*\***<br>(0.08, 0.59) | **0.10\*\*\***<br>(0.04, 0.25) |
| Very Conservative | 0.96<br>(0.29, 3.18) | **0.10\*\*\***<br>(0.03, 0.29) | **0.10\*\*\***<br>(0.04, 0.28) |
| **Self/close friend/relative diagnosed with COVID-19**<br>(referent=No) | 0.87<br>(0.46, 1.65) | 0.98<br>(0.50, 1.90) | 1.12<br>(0.66, 1.92) |
| **Not employed** (referent=Employed) | 0.96<br>(0.52, 1.75) | 1.25<br>(0.70, 2.23) | 1.31<br>(0.82, 2.08) |
| **Urbanicity** (referent=Urban) |  |  |  |
| Suburban | 0.79<br>(0.40, 1.56) | 0.67<br>(0.35, 1.27) | 0.84<br>(0.48, 1.48) |
| Rural | 0.66<br>(0.32, 1.38) | 0.54<br>(0.27, 1.06) | 0.81<br>(0.43, 1.50) |
| **Census Region** (referent=Northeast) |  |  |  |
| Midwest | 2.23<br>(0.96, 5.20) | **2.66\***<br>(1.20, 5.89) | 1.19<br>(0.64, 2.23) |
| South | 1.29<br>(0.58, 2.86) | 1.56<br>(0.73, 3.30) | 1.20<br>(0.63, 2.29) |
| West | 1.26<br>(0.51, 3.08) | 1.91<br>(0.83, 4.41) | 1.52<br>(0.76, 3.04) |

\* p<0.05, \*\* p<0.01, \*\*\* p<0.001; Source: May 2020 Associated Press-National Opinion Research Center for Public Affairs Research Poll (https://apnorc.org/projects/expectations-for-a-covid-19-vaccine/)

#### Unsure Versus No Intention

Of all three intention group comparisons, the fewest statistically significant differences are observed for unsure versus no. High school/equivalent (versus college) graduates and persons with moderate (versus very liberal) political ideology each have RRRs three times higher risk of reporting unsure versus no. Conversely, respondents less likely to report unsure versus no were those with less than $30,000 (versus ≥$100,000) household income (RRR = .33 or 67% lower risk), of African American (vs. White, non-Hispanic) racial-ethnic identity (RRR = .39 or 71% lower likelihood), and the youngest (18-29) versus oldest ≥60 years (RRR = .21 or 79% lower likelihood).

#### Yes Versus No Intention

Comparing respondents reporting yes versus no intention revealed some findings similar to those for the unsure versus no analysis—i.e. respondents from the lowest versus highest household income (RRR = .24) and of African American versus White identity (RRR = .12) had lower likelihood of being unsure—but also more extensive differences. Specifically, respondents from all three age groups spanning 18-59 (versus those ≥60 years; respective RRRs = .14, .33, .33) and espousing a somewhat or very conservative political ideology (versus very liberal; respective RRRs = .21 and .10) had lower likelihood of reporting intent to receive the vaccine. The only respondents with *higher* likelihood of endorsing yes versus no were those never married (versus married/living with a partner) and living in the Midwest (versus Northeast)—both of which had approximately 2.5 times higher likelihood of planning to receive the vaccine (respective RRRs = 2.48 and 2.66).

#### Yes Versus Unsure Intention

The comparison of yes versus unsure revealed the most extensive sociodemographic differences—most of which indicated *lower* likelihood of vaccination intent. Compared to those with a bachelor’s degree or higher, persons in all other education levels had approximately 46-66% lower likelihood of expressing yes. Regarding racial-ethnic differences, African American, Hispanic, and Other identity (versus White) respondents each had approximately 50% (respective RRRs for Hispanic and Other = .46 and .47) to 70% (African American RRR = .30) lower likelihood of intending to get vaccinated. For age, again respondents ≥60 years had, respectively, approximately 2.5 and 3.0 times higher likelihoods of endorsing intent to vaccinate compared to those ages 40-59 and 30-39. For political beliefs, respondents identifying as moderate (RRR = .24), somewhat conservative (RRR = .10), or very conservative (RRR = .10) were less like than their very liberal peers to state intent to vaccinate. Lastly, males had approximately twice the likelihood of endorsing yes than did females (RRR = 2.07).

### Differences in Reasons for Intending to Vaccinate Versus Not

Table 3 shows results comparing the *specific reasons* among persons who expressed intent to vaccinate versus those with *no vaccination intention*. Despite the conceptual and empirical variability in the specific reasons (see figure 1), the model results show considerable consistency. Specifically, the following respondents reported lower likelihood of intention to vaccinate: those with <$30,000 (versus ≥$100,000) income, African Americans (versus Whites), all age groups (versus those 60 years or older), and persons having somewhat or very conservative political beliefs. Related, Hispanic respondents were less likely to endorse chronic conditions as a reason to vaccinate versus having no vaccination intention. Conversely, respondents who were never married (versus married) and lived in the Midwest (versus Northeast) were both more likely to endorse each specific reason to vaccinate versus have no intention to vaccinate.

**Table 3:**
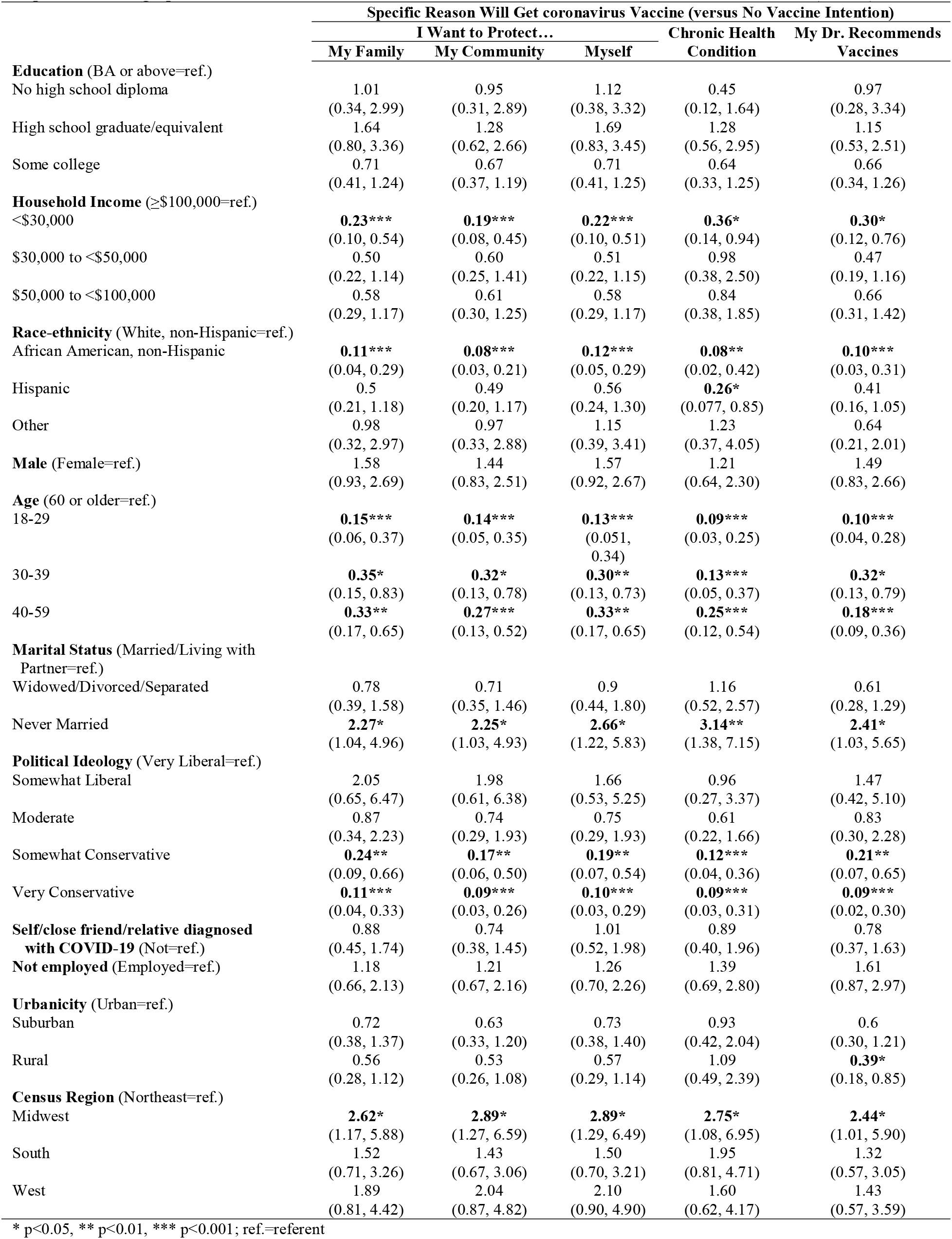
Adjusted relative risk ratios (95% confidence intervals) from multinomial logistic models estimating associations between US respondents’ demographic characteristics and intention to receive a coronavirus vaccine versus *no vaccine intention* (n = 1000).

|  | Specific Reason Will Get coronavirus Vaccine (versus No Vaccine Intention) |  |  |  |  |
| --- | --- | --- | --- | --- | --- |
|  | I Want to Protect... |  |  | Chronic Health Condition | My Dr. Recommends Vaccines |
|  | My Family | My Community | Myself |  |  |
| **Education** (BA or above=ref.) |  |  |  |  |  |
| No high school diploma | 1.01<br>(0.34, 2.99) | 0.95<br>(0.31, 2.89) | 1.12<br>(0.38, 3.32) | 0.45<br>(0.12, 1.64) | 0.97<br>(0.28, 3.34) |
| High school graduate/equivalent | 1.64<br>(0.80, 3.36) | 1.28<br>(0.62, 2.66) | 1.69<br>(0.83, 3.45) | 1.28<br>(0.56, 2.95) | 1.15<br>(0.53, 2.51) |
| Some college | 0.71<br>(0.41, 1.24) | 0.67<br>(0.37, 1.19) | 0.71<br>(0.41, 1.25) | 0.64<br>(0.33, 1.25) | 0.66<br>(0.34, 1.26) |
| **Household Income** (≥\$100,000=ref.) | | | | | |
| <\$30,000 | **0.23\*\*\***<br>(0.10, 0.54) | **0.19\*\*\***<br>(0.08, 0.45) | **0.22\*\*\***<br>(0.10, 0.51) | **0.36\***<br>(0.14, 0.94) | **0.30\***<br>(0.12, 0.76) |
| \$30,000 to <\$50,000 | 0.50<br>(0.22, 1.14) | 0.60<br>(0.25, 1.41) | 0.51<br>(0.22, 1.15) | 0.98<br>(0.38, 2.50) | 0.47<br>(0.19, 1.16) |
| \$50,000 to <\$100,000 | 0.58<br>(0.29, 1.17) | 0.61<br>(0.30, 1.25) | 0.58<br>(0.29, 1.17) | 0.84<br>(0.38, 1.85) | 0.66<br>(0.31, 1.42) |
| **Race-ethnicity** (White, non-Hispanic=ref.) |  |  |  |  |  |
| African American, non-Hispanic | **0.11\*\*\***<br>(0.04, 0.29) | **0.08\*\*\***<br>(0.03, 0.21) | **0.12\*\*\***<br>(0.05, 0.29) | **0.08\*\***<br>(0.02, 0.42) | **0.10\*\*\***<br>(0.03, 0.31) |
| Hispanic | 0.5<br>(0.21, 1.18) | 0.49<br>(0.20, 1.17) | 0.56<br>(0.24, 1.30) | **0.26\***<br>(0.077, 0.85) | 0.41<br>(0.16, 1.05) |
| Other | 0.98<br>(0.32, 2.97) | 0.97<br>(0.33, 2.88) | 1.15<br>(0.39, 3.41) | 1.23<br>(0.37, 4.05) | 0.64<br>(0.21, 2.01) |
| **Male** (Female=ref.) | 1.58<br>(0.93, 2.69) | 1.44<br>(0.83, 2.51) | 1.57<br>(0.92, 2.67) | 1.21<br>(0.64, 2.30) | 1.49<br>(0.83, 2.66) |
| **Age** (60 or older=ref.) |  |  |  |  |  |
| 18-29 | **0.15\*\*\***<br>(0.06, 0.37) | **0.14\*\*\***<br>(0.05, 0.35) | **0.13\*\*\***<br>(0.051, 0.34) | **0.09\*\*\***<br>(0.03, 0.25) | **0.10\*\*\***<br>(0.04, 0.28) |
| 30-39 | **0.35\***<br>(0.15, 0.83) | **0.32\***<br>(0.13, 0.78) | **0.30\*\***<br>(0.13, 0.73) | **0.13\*\*\***<br>(0.05, 0.37) | **0.32\***<br>(0.13, 0.79) |
| 40-59 | **0.33\*\***<br>(0.17, 0.65) | **0.27\*\*\***<br>(0.13, 0.52) | **0.33\*\***<br>(0.17, 0.65) | **0.25\*\*\***<br>(0.12, 0.54) | **0.18\*\*\***<br>(0.09, 0.36) |
| **Marital Status** (Married/Living with Partner=ref.) |  |  |  |  |  |
| Widowed/Divorced/Separated | 0.78<br>(0.39, 1.58) | 0.71<br>(0.35, 1.46) | 0.9<br>(0.44, 1.80) | 1.16<br>(0.52, 2.57) | 0.61<br>(0.28, 1.29) |
| Never Married | **2.27\***<br>(1.04, 4.96) | **2.25\***<br>(1.03, 4.93) | **2.66\***<br>(1.22, 5.83) | **3.14\*\***<br>(1.38, 7.15) | **2.41\***<br>(1.03, 5.65) |
| **Political Ideology** (Very Liberal=ref.) |  |  |  |  |  |
| Somewhat Liberal | 2.05<br>(0.65, 6.47) | 1.98<br>(0.61, 6.38) | 1.66<br>(0.53, 5.25) | 0.96<br>(0.27, 3.37) | 1.47<br>(0.42, 5.10) |
| Moderate | 0.87<br>(0.34, 2.23) | 0.74<br>(0.29, 1.93) | 0.75<br>(0.29, 1.93) | 0.61<br>(0.22, 1.66) | 0.83<br>(0.30, 2.28) |
| Somewhat Conservative | **0.24\*\***<br>(0.09, 0.66) | **0.17\*\***<br>(0.06, 0.50) | **0.19\*\***<br>(0.07, 0.54) | **0.12\*\*\***<br>(0.04, 0.36) | **0.21\*\***<br>(0.07, 0.65) |
| Very Conservative | **0.11\*\*\***<br>(0.04, 0.33) | **0.09\*\*\***<br>(0.03, 0.26) | **0.10\*\*\***<br>(0.03, 0.29) | **0.09\*\*\***<br>(0.03, 0.31) | **0.09\*\*\***<br>(0.02, 0.30) |
| **Self/close friend/relative diagnosed with COVID-19** (Not=ref.) | 0.88<br>(0.45, 1.74) | 0.74<br>(0.38, 1.45) | 1.01<br>(0.52, 1.98) | 0.89<br>(0.40, 1.96) | 0.78<br>(0.37, 1.63) |
| **Not employed** (Employed=ref.) | 1.18<br>(0.66, 2.13) | 1.21<br>(0.67, 2.16) | 1.26<br>(0.70, 2.26) | 1.39<br>(0.69, 2.80) | 1.61<br>(0.87, 2.97) |
| **Urbanicity** (Urban=ref.) |  |  |  |  |  |
| Suburban | 0.72<br>(0.38, 1.37) | 0.63<br>(0.33, 1.20) | 0.73<br>(0.38, 1.40) | 0.93<br>(0.42, 2.04) | 0.6<br>(0.30, 1.21) |
| Rural | 0.56<br>(0.28, 1.12) | 0.53<br>(0.26, 1.08) | 0.57<br>(0.29, 1.14) | 1.09<br>(0.49, 2.39) | **0.39\***<br>(0.18, 0.85) |
| **Census Region** (Northeast=ref.) |  |  |  |  |  |
| Midwest | **2.62\***<br>(1.17, 5.88) | **2.89\***<br>(1.27, 6.59) | **2.89\***<br>(1.29, 6.49) | **2.75\***<br>(1.08, 6.95) | **2.44\***<br>(1.01, 5.90) |
| South | 1.52<br>(0.71, 3.26) | 1.43<br>(0.67, 3.06) | 1.50<br>(0.70, 3.21) | 1.95<br>(0.81, 4.71) | 1.32<br>(0.57, 3.05) |
| West | 1.89<br>(0.81, 4.42) | 2.04<br>(0.87, 4.82) | 2.10<br>(0.90, 4.90) | 1.60<br>(0.62, 4.17) | 1.43<br>(0.57, 3.59) |

\* p<0.05, \*\* p<0.01, \*\*\* p<0.001; ref.=referent

### Differences in Reasons for Intending to Vaccinate Versus Uncertain Intention

Table 4 present findings comparing the specific reasons among those who expressed intention to vaccinate versus persons who indicated they were *unsure* about planning to receive a coronavirus vaccine. Again, substantial consistency is observed across the five reason-specific models, but different patterns than what was found in the abovementioned analysis for specific intent reasons versus no intent. The following groups were generally found to be associated with lower likelihood of endorsing all or most of the listed reasons for why they would vaccinate versus reporting being unsure: all educational groups (versus a bachelor’s degree or higher), all racial-ethnic groups (versus White), ages 30-59 (versus age 60 or older),

**Table 4:** Adjusted relative risk ratios (95% confidence intervals) from multinomial logistic models estimating associations between US respondents’ demographic characteristics and intention to receive a coronavirus vaccine versus *unsure vaccine intention^1^* (n = 1000).

|  | Specific Reason Will Get coronavirus Vaccine... (versus Unsure Vaccine Intention)<sup>1</sup> |  |  |  |  |
| --- | --- | --- | --- | --- | --- |
|  | I Want to Protect... |  |  | Chronic Health Condition | My Dr. Recommends Vaccines |
|  | My Family | My Community | Myself |  |  |
| **Education** (BA or above=ref.) |  |  |  |  |  |
| No high school diploma | **0.34\***<br>(0.13, 0.91) | **0.33\***<br>(0.12, 0.91) | **0.38\***<br>(0.15, 1.00) | **0.15\*\***<br>(0.04, 0.52) | 0.33<br>(0.10, 1.03) |
| High school graduate/equivalent | **0.51\***<br>(0.28, 0.96) | **0.42\*\***<br>(0.22, 0.80) | **0.53\***<br>(0.29, 0.99) | **0.40\***<br>(0.19, 0.85) | **0.36\*\***<br>(0.18, 0.73) |
| Some college | **0.51\*\***<br>(0.32, 0.82) | **0.48\*\***<br>(0.29, 0.79) | **0.51\*\***<br>(0.32, 0.83) | **0.46\***<br>(0.25, 0.85) | **0.47\***<br>(0.26, 0.84) |
| **Household Income** (≥\$100,000=ref.) | | | | | |
| <\$30,000 | 0.71<br>(0.34, 1.52) | 0.57<br>(0.26, 1.24) | 0.68<br>(0.33, 1.42) | 1.10<br>(0.44, 2.71) | 0.93<br>(0.40, 2.16) |
| \$30,000 to <\$50,000 | 0.81<br>(0.42, 1.57) | 0.96<br>(0.48, 1.91) | 0.82<br>(0.43, 1.57) | 1.59<br>(0.72, 3.48) | 0.75<br>(0.36, 1.60) |
| \$50,000 to <\$100,000 | 0.87<br>(0.51, 1.48) | 0.90<br>(0.52, 1.56) | 0.86<br>(0.51, 1.47) | 1.25<br>(0.65, 2.40) | 0.99<br>(0.53, 1.83) |
| **Race-ethnicity** (White, non-Hispanic=ref.) |  |  |  |  |  |
| African American, non-Hispanic | **0.28\*\***<br>(0.11, 0.71) | **0.21\*\***<br>(0.08, 0.54) | **0.30\*\***<br>(0.12, 0.72) | 0.22<br>(0.05, 1.04) | **0.26\***<br>(0.08, 0.79) |
| Hispanic | **0.39\*\***<br>(0.19, 0.78) | **0.38\*\***<br>(0.19, 0.78) | **0.43\***<br>(0.22, 0.85) | **0.19\*\***<br>(0.064, 0.59) | **0.32\*\***<br>(0.14, 0.71) |
| Other | **0.44\***<br>(0.20, 0.96) | **0.44\***<br>(0.20, 0.96) | 0.51<br>(0.24, 1.09) | 0.55<br>(0.21, 1.47) | **0.29\*\***<br>(0.12, 0.67) |
| **Male** (Female=ref.) | **2.04\*\***<br>(1.30, 3.22) | **1.88\*\***<br>(1.17, 3.02) | **2.04\*\***<br>(1.29, 3.20) | 1.56<br>(0.87, 2.79) | **1.92\***<br>(1.15, 3.21) |
| **Age (60 or older=ref.)** |  |  |  |  |  |
| 18-29 | 0.70<br>(0.29, 1.66) | 0.66<br>(0.27, 1.59) | 0.63<br>(0.27, 1.48) | 0.41<br>(0.15, 1.17) | 0.49<br>(0.19, 1.28) |
| 30-39 | **0.35\*\***<br>(0.17, 0.72) | **0.33\*\***<br>(0.16, 0.69) | **0.31\*\***<br>(0.15, 0.64) | **0.13\*\*\***<br>(0.053, 0.33) | **0.32\*\***<br>(0.15, 0.71) |
| 40-59 | **0.40\*\*\***<br>(0.23, 0.68) | **0.32\*\*\***<br>(0.19, 0.56) | **0.40\*\*\***<br>(0.23, 0.68) | **0.30\*\*\***<br>(0.16, 0.58) | **0.21\*\*\***<br>(0.12, 0.39) |
| **Marital Status** (Married/Living with Partner=ref.) |  |  |  |  |  |
| Widowed/Divorced/Separated | 1.36<br>(0.72, 2.57) | 1.26<br>(0.66, 2.41) | 1.54<br>(0.83, 2.86) | 2.01<br>(0.95, 4.26) | 1.04<br>(0.53, 2.06) |
| Never Married | 1.68<br>(0.88, 3.21) | 1.68<br>(0.86, 3.28) | **1.95\***<br>(1.03, 3.68) | **2.35\***<br>(1.16, 4.76) | 1.77<br>(0.84, 3.74) |
| **Political Ideology** (Very Liberal=ref.) |  |  |  |  |  |
| Somewhat Liberal | 0.58<br>(0.22, 1.53) | 0.56<br>(0.21, 1.49) | 0.48<br>(0.18, 1.29) | **0.28\***<br>(0.084, 0.91) | 0.42<br>(0.14, 1.23) |
| Moderate | **0.26\*\***<br>(0.12, 0.58) | **0.23\*\*\***<br>(0.10, 0.51) | **0.22\*\*\***<br>(0.10, 0.51) | **0.19\*\*\***<br>(0.07, 0.49) | **0.25\*\***<br>(0.10, 0.61) |
| Somewhat Conservative | **0.11\*\*\***<br>(0.045, 0.28) | **0.08\*\*\***<br>(0.03, 0.22) | **0.09\*\*\***<br>(0.04, 0.23) | **0.06\*\*\***<br>(0.02, 0.17) | **0.10\*\*\***<br>(0.04, 0.28) |
| Very Conservative | **0.11\*\*\***<br>(0.04, 0.31) | **0.09\*\*\***<br>(0.03, 0.26) | **0.10\*\*\***<br>(0.04, 0.28) | **0.09\*\*\***<br>(0.03, 0.32) | **0.09\*\*\***<br>(0.03, 0.30) |
| **Self/close friend/relative diagnosed with COVID-19** (Not=ref.) | 1.03<br>(0.59, 1.79) | 0.87<br>(0.51, 1.50) | 1.16<br>(0.68, 1.99) | 1.02<br>(0.51, 2.06) | 0.90<br>(0.48, 1.70) |
| **Not employed** (Employed=ref.) | 1.24<br>(0.77, 2.00) | 1.28<br>(0.79, 2.07) | 1.31<br>(0.82, 2.10) | 1.46<br>(0.79, 2.68) | 1.68<br>(1.00, 2.83) |
| **Urbanicity** (Urban=ref.) |  |  |  |  |  |
| Suburban | 0.91<br>(0.51, 1.60) | 0.8<br>(0.45, 1.42) | 0.91<br>(0.52, 1.61) | 1.17<br>(0.57, 2.38) | 0.76<br>(0.40, 1.42) |
| Rural | 0.84<br>(0.45, 1.59) | 0.8<br>(0.42, 1.55) | 0.85<br>(0.46, 1.60) | 1.63<br>(0.78, 3.42) | 0.59<br>(0.28, 1.20) |
| **Census Region** (Northeast=ref.) |  |  |  |  |  |
| Midwest | 1.17<br>(0.62, 2.22) | 1.29<br>(0.68, 2.46) | 1.29<br>(0.68, 2.43) | 1.23<br>(0.57, 2.65) | 1.10<br>(0.53, 2.30) |
| South | 1.18<br>(0.61, 2.27) | 1.12<br>(0.58, 2.14) | 1.16<br>(0.61, 2.23) | 1.52<br>(0.70, 3.30) | 1.02<br>(0.48, 2.16) |
| West | 1.51<br>(0.74, 3.08) | 1.63<br>(0.80, 3.31) | 1.66<br>(0.83, 3.36) | 1.26<br>(0.56, 2.86) | 1.13<br>(0.52, 2.49) |

\* p<0.05, \*\* p<0.01, \*\*\* p<0.001; ref.=referent
<sup>1</sup> Refers to Participant answering "Not Sure" when asked "If a vaccine against the coronavirus becomes available, do you plan to get it?"

## Discussion

The prospect of a safe, effective coronavirus vaccine holds promise for enabling the US to mitigate the substantial COVID-19 morbidity and mortality it has been experiencing. However, the present findings indicate that such enthusiasm for a vaccine is unequally distributed along sociodemographic lines. This analysis of a national sample reveals substantial demographic subgroup differences not only among those expressing intent versus none, but also those expressing intent versus *uncertainty*. At the time of this writing, only a few vaccine candidates have even entered Phase III trials. (19) Hence, any potential vaccine is a hypothetical consideration. However, these findings raise important concerns for what may lie ahead if a vaccine is ultimately approved by the US Food and Drug Administration (FDA) and efforts are made to design and implement effective, nation-wide vaccination campaigns.

Most broadly, they point to the challenges that lie ahead for achieving significant population uptake to ensure herd immunity. Considering the coronavirus pandemic’s substantial health, social, and economic impacts, it is troubling that only approximately 50% of respondents expressed intention to get a vaccine that would potentially protect them and others from disease and death, and, collectively, would facilitate a re-opening of society and re-engagement in many activities currently curtailed due to the risk. One may be heartened that an additional 30% reported being unsure, as that may indicate a significant number of “fence-sitters” (20) who are willing to potentially consider it and may be easier to convince than the approximately 20% who said, “No.” However, as Table 3 shows, there is a comparatively larger number of differences observed between the unsure and no groups than there are for the yes versus unsure groups. Hence, while there are some important differences between the unsure and no groups, these findings highlight how convincing unsure respondents to say “yes” will remain a challenge.

The subgroup differences also show the potential for widening health disparities. Extensive attention has been paid to the disproportionate COVID-19 exposure risks and morbidity and mortality burdens that non-White and lower income subpopulations have experienced to date. (10,11) Yet, the present findings suggest that the opportunity to substantially mitigate such disparate risks via an effective vaccine will be unequally realized—notably by racial-ethnic and SES groups already significantly impacted by COVID-19—with the troubling potential for current disparities widening.

The findings on specific intentions to vaccinate are helpful in this regard. High prevalence was observed across three of the five specific reasons that, altogether, lend themselves well to health promotion efforts. Nevertheless, having one’s doctor recommend vaccines had the lowest endorsement, which is concerning as health care provider expertise is often a focus in addressing vaccine hesitancy. Also, despite the variation in support across these five items, they still produced very similar patterns of findings in the multivariable models. This suggests that it is important to start considering a range of local, county, state, and federal strategies for outreach, trust building, and education/promotion to marginalized and other demographics. Vaccine mandates are not a simple answer to uptake disparities—they can be politically fraught (especially in the current US climate where science has been heavily politicized) and counter-productive to the goals of maximizing vaccine uptake. As such, they should be viewed as a last resort. (21) Furthermore, it is important to consider how such hesitancy attitudes may exist due to context. For example, expressing uncertain or no intention to receive the vaccine may reflect previous or current barriers to obtaining preventive health care (i.e. people reporting little to no intention to get the vaccine due to a belief that it will be difficult to obtain) and distrust of medical professionals and/or the vaccine itself. (22)

## Limitations

Several study limitations should be acknowledged. First, these data were collected in mid-May 2020. Thus, responses could have been affected by specific media coverage at that time period about vaccine development or Operation Warp Speed, the severity of cases and deaths, and extent of re-opening policies throughout the nation.

Second, this analysis only focused on adults. Vaccination will also be important for children in order to ensure their safety and achieve herd immunity for others. However, parents’ attitudes towards having their children receive a COVID-19 vaccine are unclear and require more research.

Third, respondents were asked about receiving a hypothetically available vaccine. It is possible that people may have different attitudes if a vaccine is eventually made available, as it will have a reported safety and effectiveness profile.

Fourth, while the analysis focused on different reasons for intentions to receive the vaccine, it did not examine why certain respondents stated they either had no intention to receive the vaccine or were unsure. This was due to survey design and data limitations. The survey included a series of nine items asked of those respondents who initially stated no intention to receive the vaccine. Yet, due to the small proportion of such respondents in the overall sample and the small number of endorsements for each specific reason (all but one reason endorsed by less than 69 respondents), it was not possible to do a systematic modeling of subgroup comparisons of those responses. This is instructive for other studies in that comprehensive analyses of hesitancy will require (a) sample sizes larger than the 1000 respondents in this analysis and (b) specific reason items for those answering unsure as well as yes and no.

## Implications for Future Research

Overall, these findings offer useful insights—particularly with respect to three valid categories of intention: no, unsure, and yes. First, future surveys need to include an “unsure” category, as efforts to promote vaccination among the uncertain will likely need to be very different in approach and design than efforts aimed at those who have no intention of getting vaccinated.

Second, in terms of measurement, it is important to be mindful that a “yes” response in this or any future survey is capturing *potential* to receive the vaccine (under hypothetical circumstances) and is just one of multiple factors that a person’s receipt is contingent upon. Hence, such “yes” estimates should not be taken for granted. How such intentions may fare if a vaccine is released will depend on access issues that may facilitate or hinder receipt. Translating such willingness into actual effort to receive the vaccine will require designing programs that offer the public sufficient information about its safety and effectiveness and make its receipt as convenient as possible. Likewise, per research on diffusion of innovation (23), heterogeneity likely exists among the yes and uncertain responses in willingness to receive the vaccine. Though people may express willingness to receive this new innovation, they may not necessarily intend to be the “innovators” and “early adopters” (i.e. first in line), but rather wait for others to receive it first to observe from people in their networks what the side effects might be like.

Third and finally, COVID-19 vaccine-related attitudes and receipt intentions need to be assessed repeatedly by national polls. Public health efforts would greatly benefit from survey firms and polling organizations making their data easily available to the public health community (like NORC has) to conduct more extensive analyses than what are typically presented in polling organization’s public audience- and news media-focused reports. At a time when significant amounts of COVID-19-related disinformation are circulating and science is being heavily politicized and challenged amidst the tremendous burdens of the pandemic itself, quality information on the public’s vaccine intentions and the reasons for such intentions are crucial— especially as multiple Phase III trials proceed and the design of effective vaccination campaigns nation-wide becomes increasingly more urgent.

## Data Availability

The data for this study are publicly available on the NORC website and were accessed by the study author via https://apnorc.org/projects/expectations-for-a-covid-19-vaccine/

## Acknowledgements

RMC received no funding for conducting this study. Thanks extended to Peter Hotez for his suggestions on this paper.

## Abbreviations

AP: Associated Press
FDA: Food and Drug Administration
RRR: relative risk ratio
SES: Socioeconomic status

